# Estimative of real number of infections by COVID-19 on Brazil and possible scenarios

**DOI:** 10.1101/2020.05.03.20052779

**Authors:** H. P. C Cintra, F. N. Fontinele

**Affiliations:** Physics Institute, University of Brasilia, Brasilia, Brazil

## Abstract

This paper attempts to provide methods to estimate the real scenario of the novel coronavirus pandemic crisis on Brazil and the states of Sao Paulo, Pernambuco, Espirito Santo, Amazonas and Distrito Federal. By the use of a SEIRD mathematical model with age division, we predict the infection and death curve, stating the peak date for Brazil and these states. We also carry out a prediction for the ICU demand on these states for a visualization of the size of a possible collapse on the local health system. By the end, we establish some future scenarios including the stopping of social isolation and the introduction of vaccines and efficient medicine against the virus.

## I. INTRODUCTION

On December 2019, the city of Wuhan on mainland China started experiencing an outbreak of unknown pneumonia cases. Later, the cause of this outbreak was identified as a virus belonging to the *Orthocoronavidae* subfamiliy and the *Betacoronavirus* genus [1], similar to the SARS-CoV virus that caused the SARS crisis on 2003 [2]. That similarity suggested the name SARS-CoV-2 to the novel coronavirus, and COVID-19 to the disease.

The virus quickly spread to other countries, reaching several countries by the end of February and being declared as a pandemic crisis by the World Health Organization (WHO) at 11th March, being classified as a threat of high risk for the world [3]. Since then, several mathematical models were used to predict the dynamics of the pandemic crisis on other countries. One of those models with the biggest impact was developed by the Imperial College London [4].

On Brazil, the first case registered dates back to 25th February, but on this study we suggest evidence that the infection might have started 19 to 24 days before the official record. We then proceed to simulate the crisis on specific states and attempt to estimate the real scale of the outbreak, predicting when the infections peak might occur as well as the curve for ICU demand for each of those states. Finally, we present some future scenarios based on how the stop of the intervention might affect the curve and how the introduction of vaccines or available medicine might also change the infection curve since there are several studies being made to evaluate possible use of pharmaceutical drugs to cure the disease [5], [6] and [7].

## II. MODEL DESCRIPTION

We make use of a SEIRD model, dividing the population into 5 groups: Susceptible, Exposed, Infected, Recovered and Dead. The exposed population differs from the infected population on the development of symptoms; an individual with the virus enters first the exposed group, carrying the virus during the incubation period; then, with the development of symptoms, the individual passes to the infected group. The rate of infection is proportional to the number of infected and a contact constant *β* given by the average number of contacts between individuals times the probability of contracting the virus on each contact. The rate of symptoms development is proportional to the incubation period *c*^−1^. The rate of recovery is proportional to the percentage of people who recovers divided by the average time taken from symptoms onset to recovery, similarly to the death rate. Another consideration is that people on the exposed group might infect susceptible people with an infection rate k which is a small fraction of *β*.

The following diagram represents the dynamics of these populations:

**Fig. 1:**
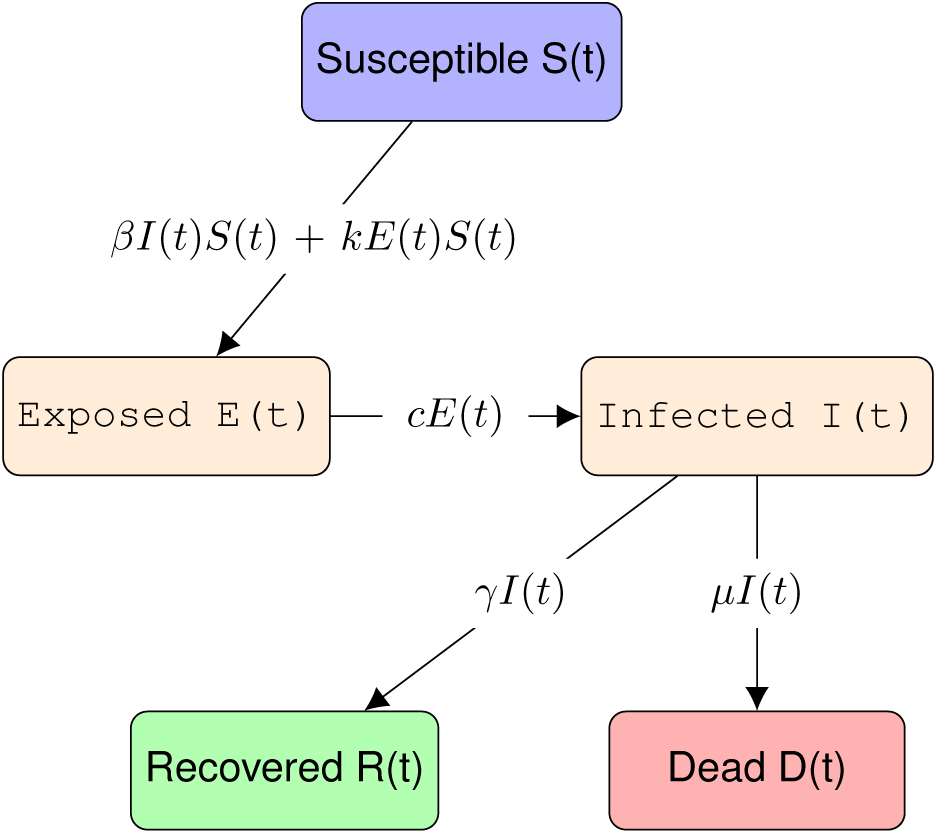
Representation of a SEIRD model, a susceptible person gets exposed to the virus, being infected afterwards and either dies or recovers from the disease.

This model is represented by the following set of differential equations

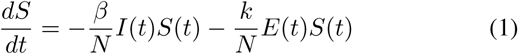

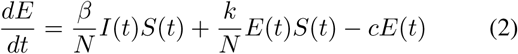

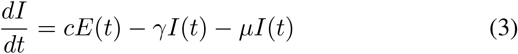

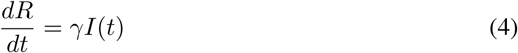

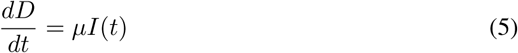

where the recovery rate *γ* and death rate *μ* are represented in terms of the Infection Fatality Rate (IFR) *P*_:(_ and the average time from symptoms onset to recovery *τ_r_* and death *τ_d_*.

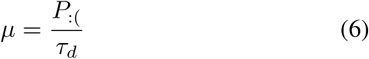

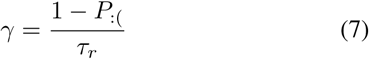

All equations described conserve the total population *N*, which is assumed constant and homogeneous for the model to be valid. This, of course, presents a limitation of the model, since in reality *N* is not homogeneous. Therefore, here *N* carries the role of effective population, being equivalent to the population in which the virus might get to under the interval of some months. Estimating the real *N* is not an easy task, on next sessions we discuss how we decided to estimate this number.

We then, divided the population into age groups to better describe how these rates vary from group to group. With that, we suggest the following changes already proposed by [8]:

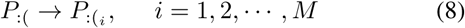

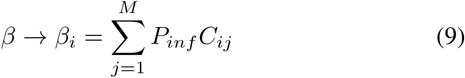

where *M* is the number of age groups, *C_ij_* is the social contact matrix, representing the average contacts between a member of the *i*-th group with all other *j*-th groups and *P_inf_* is the probability of being infected at each contact.

With these definitions, we represent non-pharmaceutical interventions such as social isolation and lock-down with a decrease of *β* given by a logistic function of the type

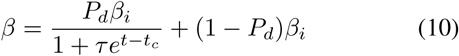

here, *β_i_* is the infection rate before the intervention, *t_c_* is the time when the intervention starts, *P_d_* is the fraction of reduction achieved and *τ* is a constant related to the time taken from the start of the intervention until *P_d_* is reached.

When simulating the curve for infections and deaths from Brazil and the states of Pernambuco, Espirito Santo, Distrito Federal, Sao Paulo and Amazonas, we used the model described above. Meanwhile, when simulating the ICU demand, we do not apply the age division for lack of specific data for each age group, thus, we apply the simple SEIRD model with *β* extracted from the fitting of data of each state and *P*_:(_, *τ_r_* and *τ_d_* appropriate for ICU patients by COVID-19.

## III. ESTIMATING THE PERCENTAGE OF LOST CASES

### A. Number of hospitalizations by SARS

According to [9], 3.6% of COVID-19 infections are severe and require hospitalization and 30% of those critical and require an ICU unit. Some studies found an hospitalization rate around 14% [10], another study found similar percentages, stating that 19% of the infections resulted on hospitalizations [11]. However, these studies take these fractions according to the registered cases, which is undernotified in many regions.

With the emergence of the novel coronavirus, the number of hospitalizations by SARS per week increased when compared to the years of 2019, 2018 and 2017. Using the number of hospitalizations by SARS on those years, we construct an background behavior, that is, the expected number of hospitalizations due to other respiratory diseases (Figure 2). The number released by the Health Ministry per week is subjected to alterations due to new results on the following weeks regarding the one released. For example, by the end of the 6th week of the year 2018, the official report estimates a number of hospitalizations around 50, but on later on, this number was corrected to be close to 200. Because of this uncertainty on the most recent data, for this estimation we use the values available a few weeks before the most recent released (Figure 3).

**Fig. 2:**
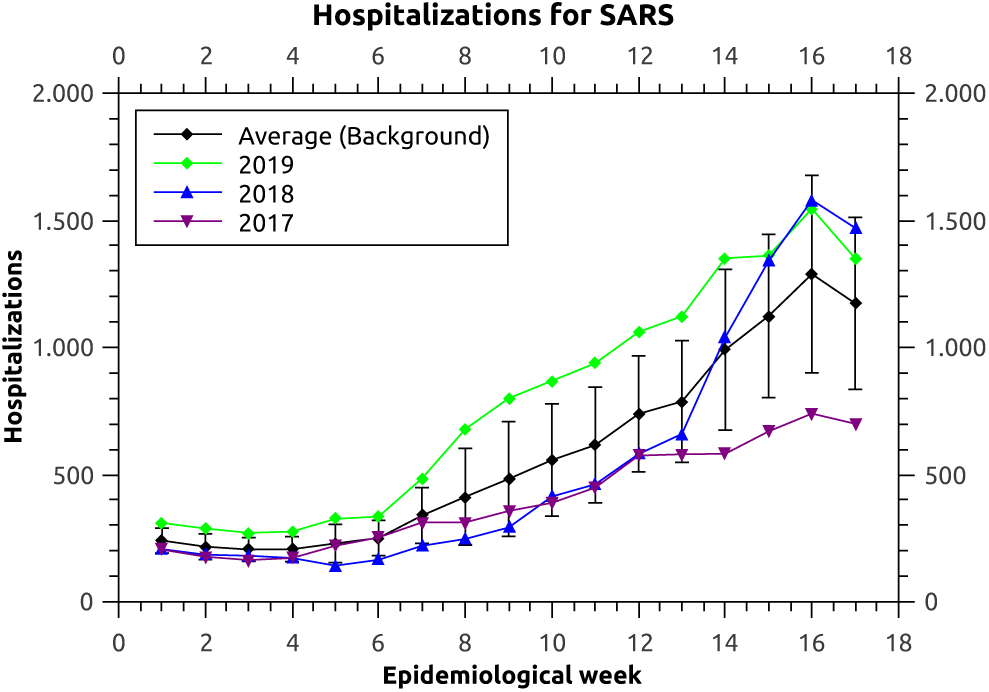
Hospitalizations by SARS on Brazil on the years of 2017, 2018 and 2019.

**Fig. 3:**
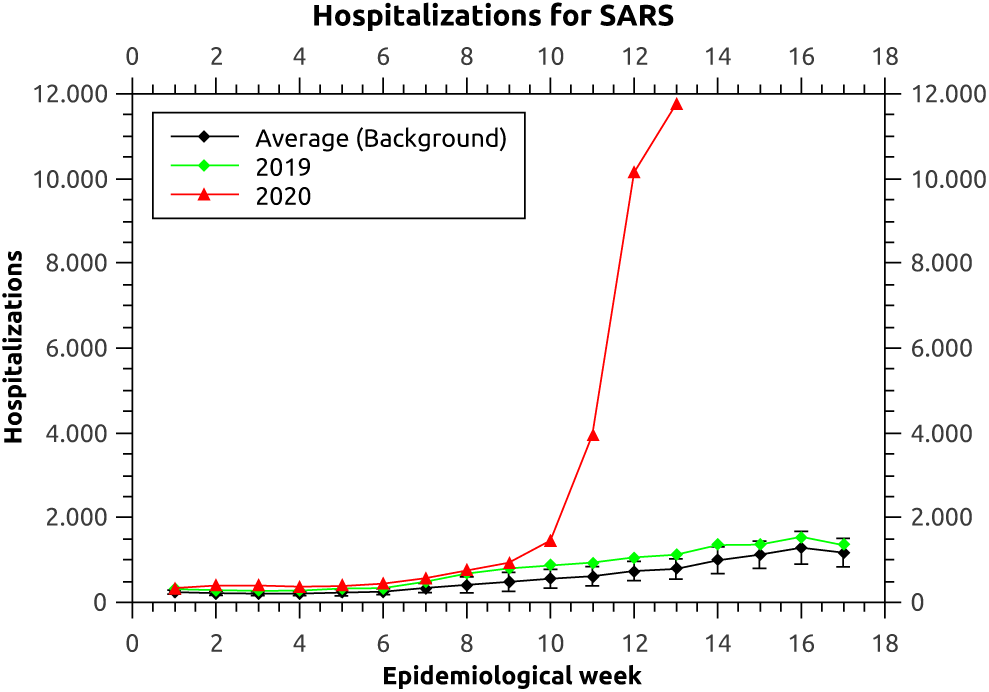
Number of hospitalizations by SARS on Brazil on the year of 2020, 2019 and the background average.

From the data, at the 6th week, the number of hospitalizations was higher than the upper error bar of the background by 121 hospitalizations, and higher than the year of 2019 by 106, an increase of 31%. According to a study done on COVID-19 patients on Shanghai, the hospitalization occurs on average 4 days after symptoms onset, ranging from 2 to 7 [12], this study, together with the increase of SARS hospitalizations by the 6th week of 2020 suggests the possible existence of COVID-19 cases on Brazil around February 1st to February 6th, 19 to 24 days before the official record of the first case on 25th February.

Following the increase of cases, by the end of the 13th epidemiological week of 2020 (28/03/2020), the number of hospitalizations by SARS on Brazil was already, 12260, while the upper error bar of the background is 1028, and the year of 2019 registered 1123. Supposing that, 90% of the excess of hospitalizations is due to COVID-19; based on the observation that the year of 2019 is about 10% bigger than the background, meaning we could see this behavior on 2020 as well; that marks around 10023 to 10108 hospitalizations by infections of the SARS-CoV-2 virus, which reflects on 278416 to 280777 infections between 21/03/2020 and 26/03/2020 (According to the average time taken to hospitalization). The comparison with the official numbers reported on this period gives a real number 90 to 200 times bigger than the one released (24, using the average). That represents a lost of 99.2% (99.0 - 99.5) of the infections. By comparison, a study done on China found 86% of undocumented infections prior to 23th january [13].

### B. Number of tests performed

A study of the Imperial College London estimated the number of infections on 11 European countries until 28th March, based on the basic reproduction number of the disease, found to be between 2 and 3 [14], [15], [16] and [17], and the type of non-pharmaceutical intervention done by the countries on specific dates [18]. With these estimations, we may find the percentage of lost cases on these countries until 28th March by comparing the estimate number of people infected with the official data available at 28th March. Relating these percentages with the number of tests done per 1000 habitants and the number of tests done per day per 1000 habitants, we got an linear relation between the number of total tests done per 1000 habitants and per day per 1000 habitants on a country and the percentage of lost cases (Figure 4 and 5).

**Fig. 4:**
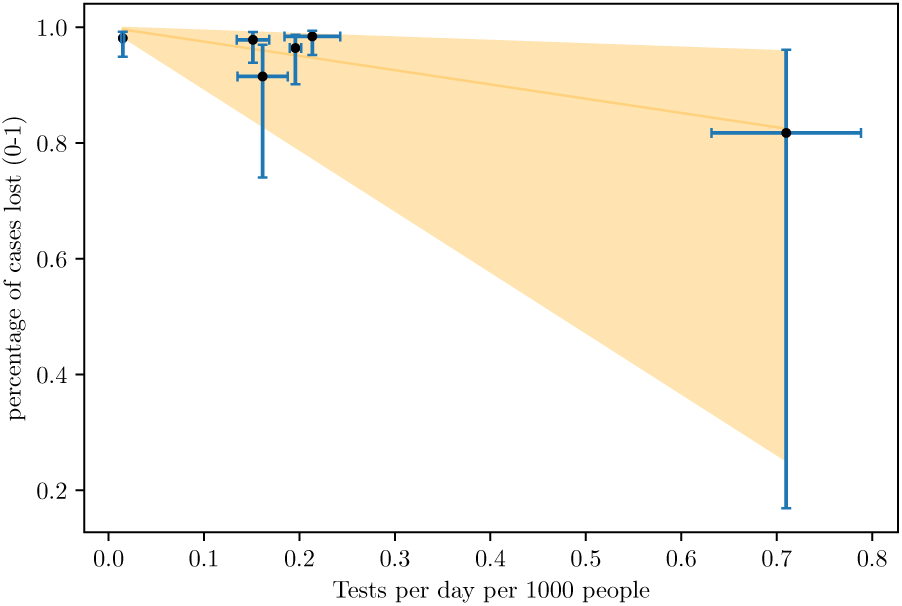
Relation between the number of tests performed per day per 1000 habitants and the fraction of lost cases.

**Fig. 5:**
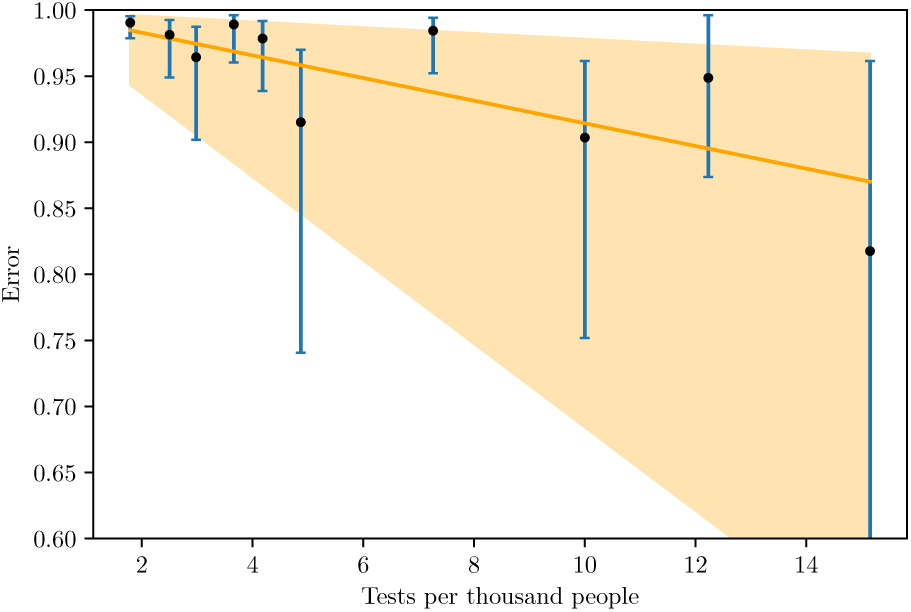
Relation between the number of tests performed on total per 1000 habitants and the fraction of lost cases.

The number of points on each graph is different because, although the study considered 11 countries, not all of them had data of tests per day available on [19]. We also compared the undocumented cases with the progression of the outbreak on each country and the day on which the non-pharmaceutical interventions were imposed, but found no correlation. We evaluated the effect of the increasing rate of testing as well, but it had no observable effect. From this relation, a country needs to perform 4 (0.94 - 17) tests per day per 1000 habitants in order to obtain a excellent track of infections. Here, the large margin for the higher values of testing arises from the low density of data points on the bigger values of the x-axis on figure 4.

The last official register of the total number of tests done per 1000 habitants on Brazil was 3.46, which corresponds to 97% of cases lost (95.6 - 99.7). A more precise number could be achieved with the data of tests per day per 1000 habitants, allowing a 2-dimensional regression, unfortunately, we found no record of this information, still, when fitting the data to a 2-dimensional regression algorithm, the resulting function states that the most important factor controlling the uncertainty of cases is tests per day per 1000 habitants. That could also be observed by looking at the graphs individually, the number of total tests performed per 1000 habitants decreases the percentage of undocumented infections in a much lower rate than the number of tests per day per 1000 habitants.

Both methods found an region of agreement (99.5% to 99.7%) of undocumented infections on Brazil. With the agreement of both methods, we decided to accept the estimate for undocumented infections on Brazil and moved on to the simulations of the country and some specific regions.

## IV. SIMULATIONS

For the simulation of Brazil, we used the *World Population Prospects* from the United Nations (UN) to evaluate the age distribution on Brazil on the year of 2020 [20]. We found no study measuring the social contact matrix for the country, but the study [21] evaluated the high levels of social contact on Brazil as an important factor for the spreading of leprosy. Therefore, we decided to use the social contact matrix found with the highest entries among those available (Poland) due to Brazilian culture of proximity.

For the values of *γ* and *μ* we choose to use the ones found on South Korea, Germany, Iceland and Taiwan data, since these countries are performing more tests per 1000 habitants than Brazil, making their data more reliable (Figures 6 and 7). For each country, we acquired the average values for *τ_d_* and *τ_r_*, knowing the CFR.

**Fig. 6:**
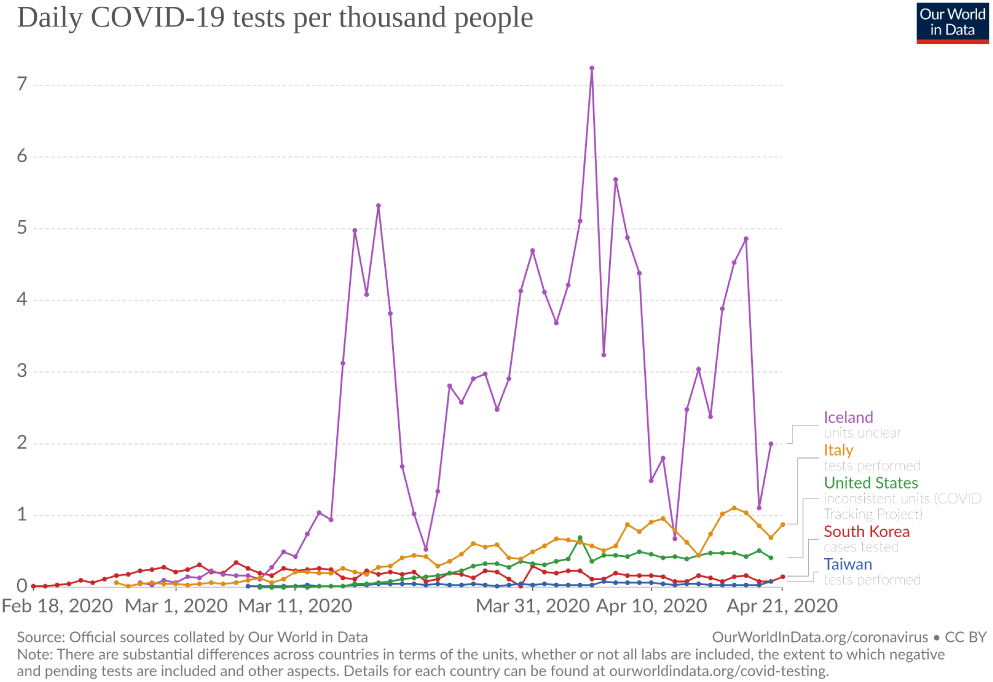
Tests Per Day Per 1000 habitants. Taken from [19].

**Fig. 7:**
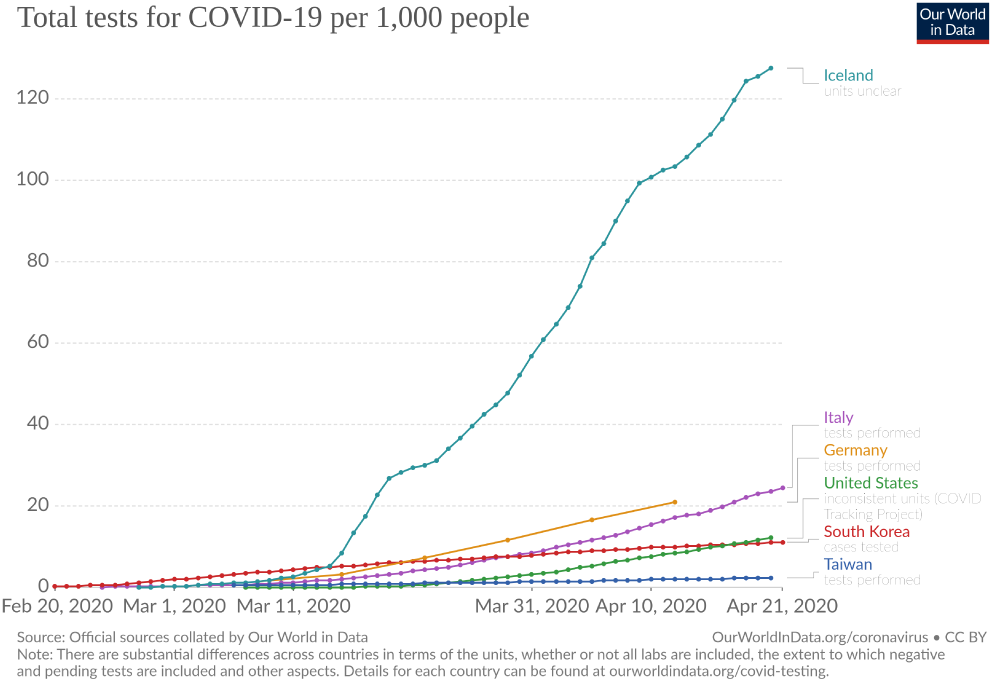
Total number of tests per 1000 habitants. Graph taken from [19].

**Table I:** Average values of *τ_d_* and *τ_r_* acquired from data.

| Country | $\tau_d$ | $\tau_r$ |
| --- | --- | --- |
| Iceland | $14.3 \pm 4.3$ days | $11.5 \pm 4.5$ days |
| South Korea | $13.5 \pm 5.6$ days | $21 \pm 10$ days |
| Germany | $13.6 \pm 5.8$ days | $17.5 \pm 8$ days |
| Taiwan | — | — |
| Average | $13.8 \pm 5.2$ days | $16 \pm 7.5$ days |

Data from Taiwan presented large fluctuations on the behavior of *μ* and *γ*, even with a almost constant Case Fatality Rate (CFR) 1.3% ± 0.2%, making the values for *τ_d_* and *τ_r_* inconclusive. That might be explained by the early intervention made by the local government, drastically changing the values for the parameters. Clinical studies performed on Wuhan patients found *τ_d_* on average 18 days (6-32) [22], and 20 days (17-24) [23].

When fitting the data of those countries with the model to extract *β* (Table II), we took into consideration on the simulations the non-pharmaceutical intervention on each country in order to better describe *β*. The value of *β* was used to set an reference to compare with the ones found with the fitting of data from each state.

**Table II:** Values of *β*.

| Country | $\beta$ |
| --- | --- |
| Taiwan | $0.427 \pm 0.066$ |
| Germany | $0.483 \pm 0.025$ |
| South Korea | $0.534 \pm 0.040$ |
| Iceland | $0.685 \pm 0.121$ |
| Average | $0.532 \pm 0.063$ |

For the incubation period *c*^−1^, we took an average found of previous studies (Table III)

**Table III:** Incubation time of the disease according to other studies with an average of 5.1 days.

| incubation time | 95% confidence | Reference |
| --- | --- | --- |
| 6.4 days | 5.6-7.7 | [24] |
| 5.2 days | 4.1-7 | [25] |
| 5 days | – | [26] |
| 4 days | – | [27] |
| 5.1 days | 4.5 - 5.8 | [28] |

The value for *k* was set to 44% of *β* based on the findings that presyntomatic cases were responsible for 44% of the infections [?]. The parameter *P*_:(_ for each age group was set re-scaling to the international average of case fatality rate (CFR) [29] with the estimated infection fatality rate of 0.7% [9], ranging from 0.001% for those younger than 20 years of age to 10.1% to those older than 80 years of age (Table IV), while *P*_:)_ = 1 − *P*_:(_.

**Table IV:** CFR of COVID-19 for different ages.

| Age (years) | Case Fatality Rate |
| --- | --- |
| 0-9 | 0.001 |
| 10-19 | 0.001% |
| 20-29 | 0.001% |
| 30-39 | 0.06% |
| 40-49 | 0.12% |
| 50-59 | 1.2% |
| 60-69 | 2.5% |
| 70-79 | 7.0% |
| +80 | 10.1% |

To simulate the ICU population, we added the new population on equations (1) to (5), the hospitalized population *H*. The introduction of this compartment is done by removing individuals from (3) with rate *P_h_/τ_h_*, where *P_h_* is the fraction of infections that are critical and require ICU units, and *τ_h_* is the average time from symptoms onset to admission on ICU. Inside the H compartment, individuals are removed to the death compartment with rate μ_h_ = *P_dh_/τ_dh_*, where *P_dh_* is the probability of dying upon ICU entry and *τ_dh_* is the average time from ICU admittance to death. Similarly, individuals are also removed to the recovered group with analogous rates *γ_h_* = (1 – *P_dh_*)/*τ_rh_*. The result is the following modification on equations (3) to (5)

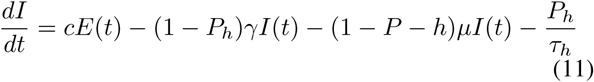

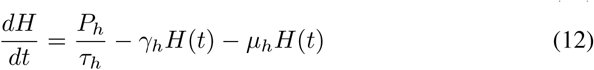

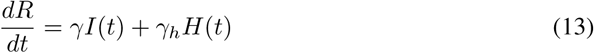

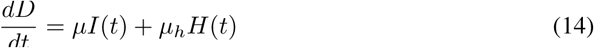

Table V contains the parameters for the simulation of ICU population.

**Table V:** Paramters for the simulation of ICU population

| Parameter | Value | Reference |
| --- | --- | --- |
| $P_h$ | 0.015 | adapted from [9] |
| $\tau_h$ | 3.5 days | [30] |
| $\tau_{rh}$ | $16 \pm 4$ days | [12] |
| $\tau_{dh}$ | 7 (3-11) days | [31] |
| $P_{dh}$ | 0.52 | [30] |

## V. RESULTS

On the simulation for the whole country, we considered *N* as 5% of the total population based on an international behavior for the total number of infections on other countries[9]. We also selected *P_inf_ =* 14% according to [8].

In order to input on the simulation, the effect of the use of masks by a large number of individuals we use a logistic function to decrease the value of *P_infec_* on 50% based on [32], the slope of the decreasing region was set to be 10x slower than the one simulated for the social distancing. We also choose *P_d_* = 0.5, taking the national average for the population on social isolation [33].

**Fig. 8:**
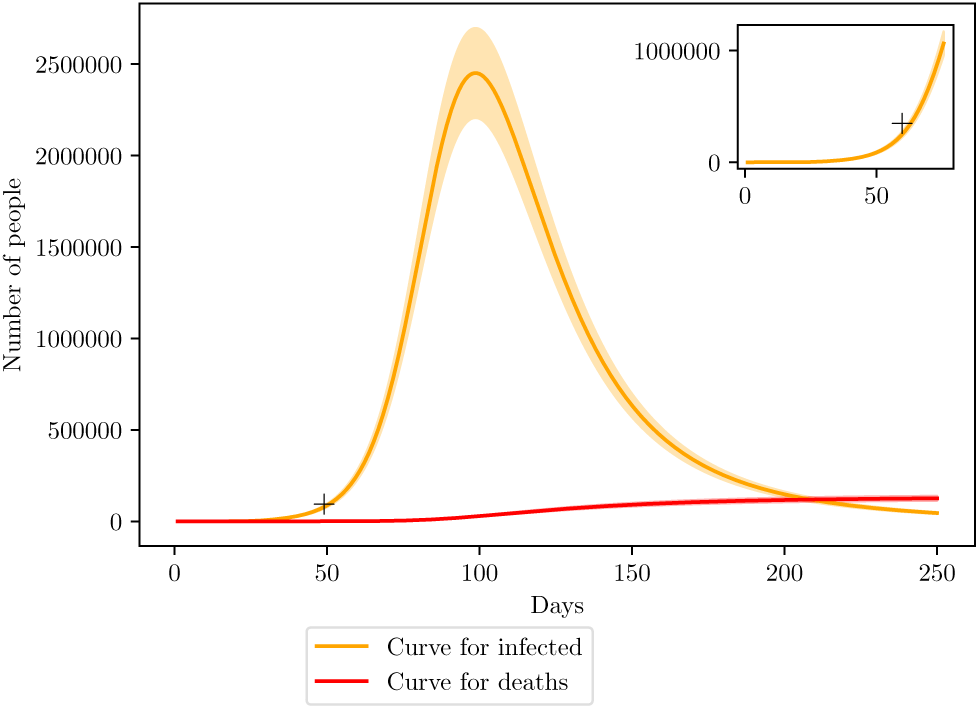
Simulation of the COVID-19 pandemic crisis on Brazil.

The curve shows a good agreement with the estimated values by the number of SARS hospitalizations on the last weeks of March, shown by the + mark on the graph. We also predict that the peak of the infection curve on Brazil should be 100 days after the first case, in which we considered to be the beginning of February. Therefore, the peak should be on the middle to end of May with a million of infections, ranging from 2.2 to 2.7 million. The number of deaths is estimated to be around 126 thousand, ranging from 114 to 139 thousand.

The shading areas represent a 10% deviation from the simulated curve. The high value of deviation was chosen as a reflection of the uncertainty on the value for effective population *N*.

### A. Pernambuco

Online data available from the local government on [34] states a total of 0.84 tests per 1000 habitants and an average of 0.05 tests per day per 1000 habitants, placing it on more than 90% of infections being undocumented. For the simulation, we acquired data regarding the age and geographical distribution of the population from the last census from IBGE [35].

The official record for the first case dates to 12th March, however, data from [34] now shows a ICU entry of a 71 year old man on the capital Recife, diagnosed with the virus SARS-CoV-2. The patient started with the symptoms on March 1st. We choose to set this date as the starting point of the simulation. According to [33], the isolation index, which measures the fraction of the population in social isolation is on average 50%.

**Fig. 9:**
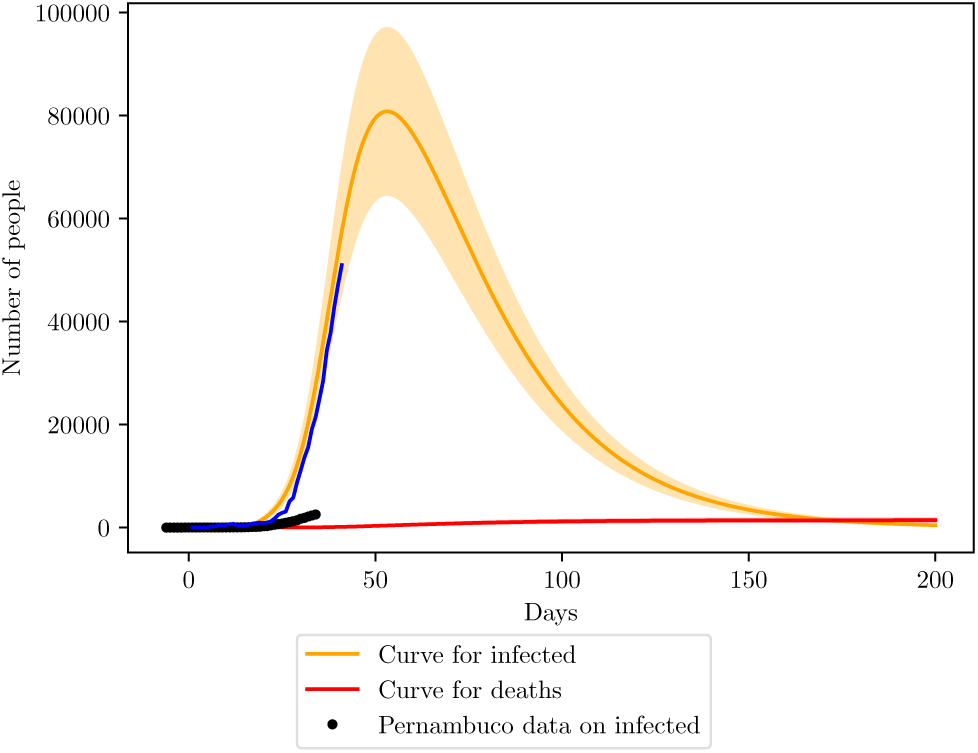
Simulation of the COVID-19 pandemic crisis on Pernambuco. The blue curve shows the behavior of the data considering 90% loss of infections.

The simulation shows a peak close to the 50th day, on the beginning of May, with 8000 infections, ranging from 6000 to 10000. The number of deaths estimated is 1400 (1167 - 1680). Here we increased the margin of error to 20%, to represent a larger uncertainty on *N* at specific locations.

Despite the large number of cases lost, when fitting the data with a simulated curve, the value of *β* is 0.460 ± 0.050, which agrees to international standards. That indicates a good tracking of the rate of change of the infection curve on Pernambuco. The state might not have the precise values of the real infections, but it has a good knowledge of their growth. That is an important feature for the state to be able to say that it’s data might represent a small scale of the real scenario.

The state of Pernambuco has a total of 1315 ICU beds according to a census carried by the Brazilian Association of Intensive Medicine (AMIB) on the year 2016 [36]. However, recent news point to 80% of these beds already being occupied, bringing the available number of ICU beds to 263.

**Fig. 10:**
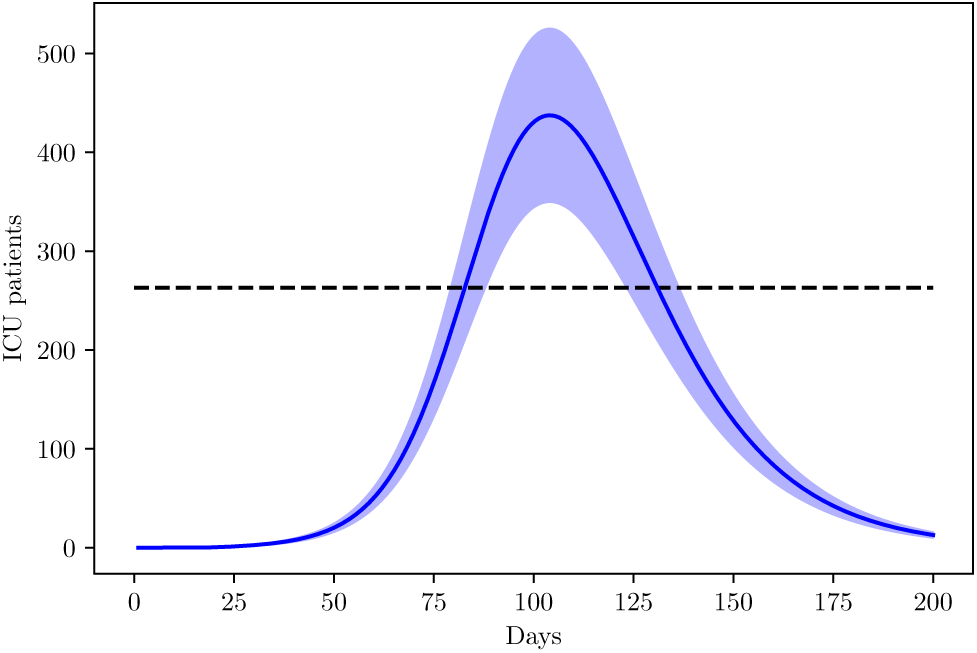
Simulation of the ICU population on Pernambuco due to COVID-19.

### B. Espirito Santo

On Espirito Santo, the online data provided by the government states a total of 6.70 tests per 1000 habitants realized, placing the uncertainty percentage close to 88% (78 – 98). There are also 161 ICU units available for COVID-19 cases [37]. The population data for the simulations was retrieved from a local census done by IBGE [38]. The isolation index is on average 45% [33].

Like Pernambuco, the fitting of Espirito Santo data reveals a good agreement of *β* with international parameters, *β* = 0.436 ± 0.199, however, the large margin of error shows low confidence on that data.

We found no record of previous hospitalizations due to COVID-19 prior the first case announced on 6th March, like the one found on Pernambuco, therefore, we choose the official day as the starting point of the disease. The first infectious individual was on the age group between 30-39 years.

**Fig. 11:**
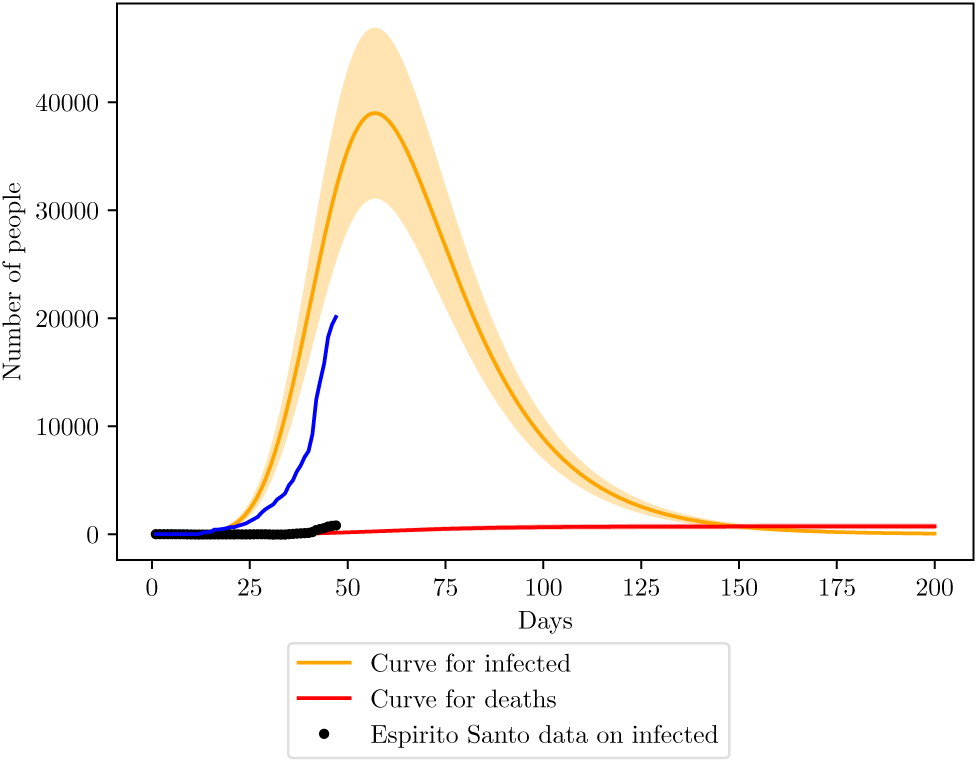
Simulation of the COVID-19 pandemic crisis on Espirito Santo.

**Fig. 12:**
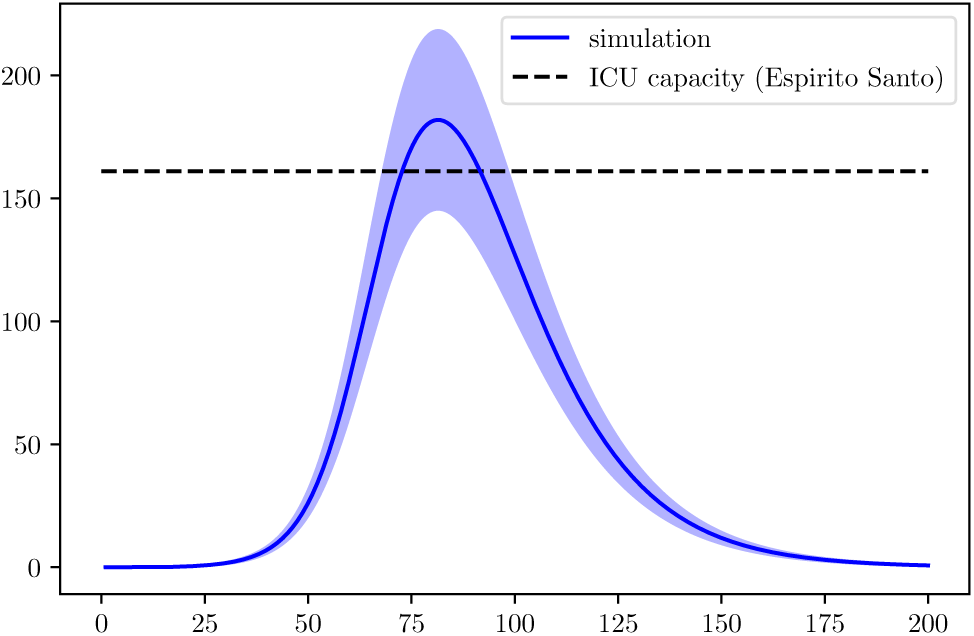
Simulation of the ICU population on Espirito Santo due to COVID-19. Blue curve represents data following the 88% of undocumented infections.

The peak of Espirito Santo is close to 70 days after the start, being this date close to 15th May, with a maximum infection number around 40000 (48000 - 32000). The number of deaths is estimated to 700 (560-840).

### C. Distrito Federal

Recent data from the government reveals 20716 tests, meaning 6.8 tests per 1000 habitants, placing the state on most likely 86% (78.5 - 94.2) of undocumented infections, unfortunately, no record of tests per day was found, so a better accuracy of lost cases was not possible. The first register of COVID-19 on the state is from 5th March, with non-pharmaceutical interventions starting at 10th March [39].

Like previous states, the IBGE census was used to extract population distribution [40].

The fit of data with the simulations returns an efficiency of 88% of the social isolation, but *β* and *τ_d_* are off the margin of acceptance, indicating that the state is not tracking well the rate of increase of deaths and cases, possibly invalidating the estimate percentage of efficiency of the social isolation. The isolation index according to [33] is on average 50%.

**Fig. 13:**
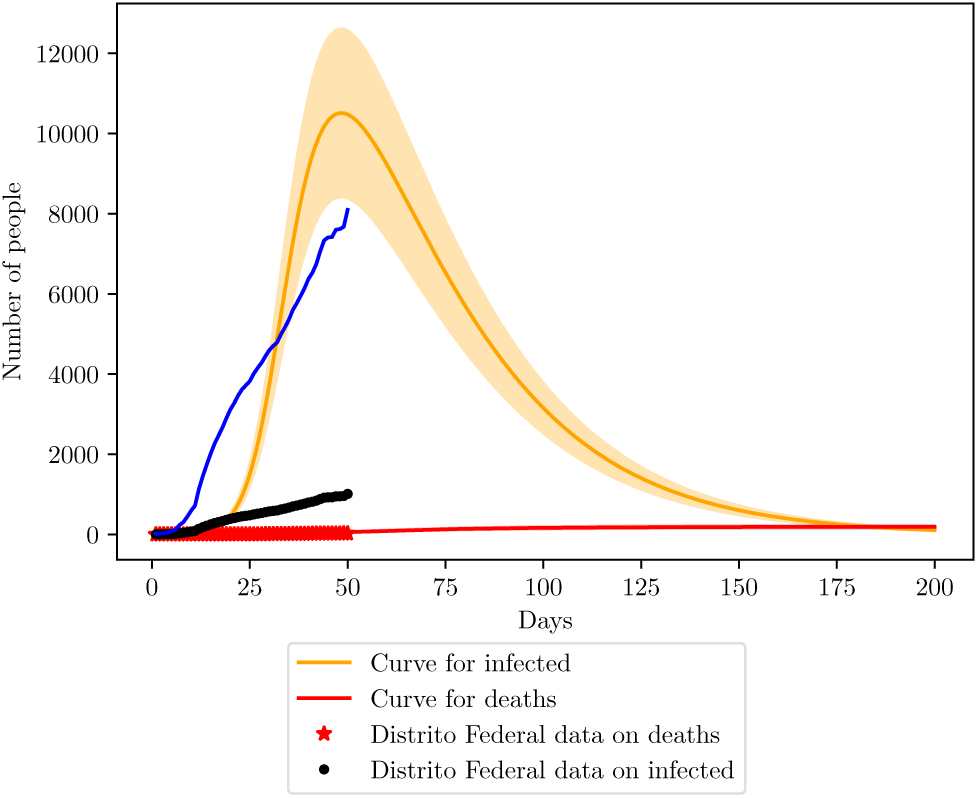
COVID-19 scenario for Distrito Federal. Blue curve represents the behavior of data considering 86% loss of infections.

The simulation shows that the Distrito Federal is currently at it’s higher number of infections, around 10000 (8000 - 12000). The maximum number of deaths is projected to 190 (158 - 228). Also, with the current number of infections, Distrito Federal is losing 89% of cases (87 - 91), in agreement with the margin estimated by the number of tests performed.

From the AMIB census, the state posses 659 ICU beds, we assume 80% of occupation before the disease reached the state.

**Fig. 14:**
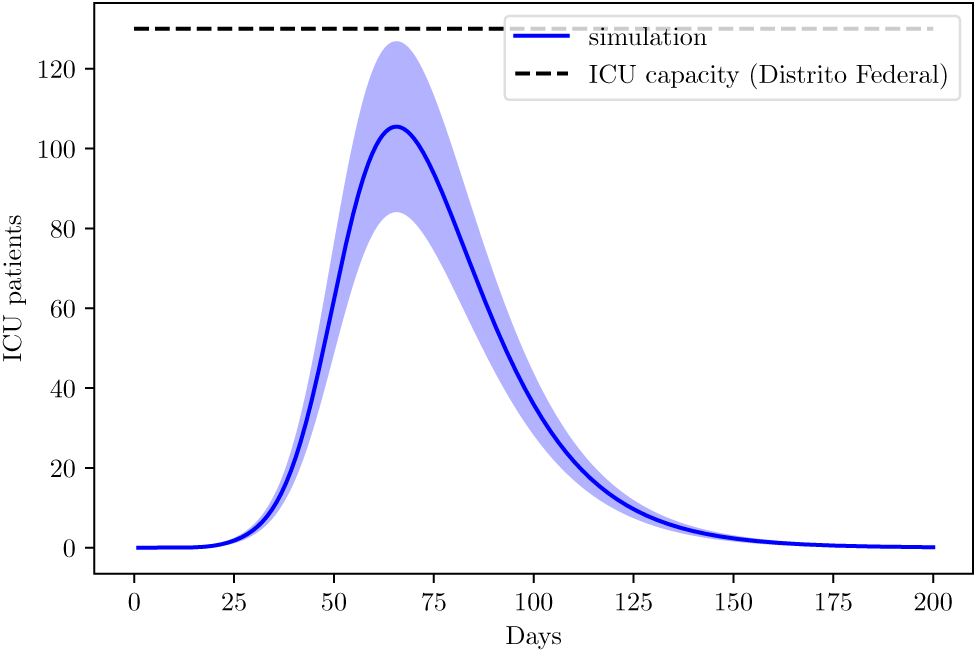
ICU demand on Distrito Federal due to COVID-19.

### D. Sao Paulo

The state of Sao Paulo also provided online data gathered by the government [41]. The first infection notified dates from 26th February. Studies done with cellphone data from Sao Paulo habitants saw an average of 53.6% ± 3.4% of the population is respecting the social isolation imposed by the local government on 24th March [41].

When fitting the data with the model, considering a non-pharmaceutical intervention starting 27 days after the first case, we found an quarantine efficiency of 58.3% ± 7%, with agreement of the study. We also found *β* = 0.454 ± 0.52, indicating that Sao Paulo is also on good track of the increasing rate of the outbreak. Unfortunately, the government did not display data on infections, but with such a high mortality, around 8%, the number of infections is probably 4x bigger than the official number (meaning 75% of undocumented infections), assuming that the number of deaths is in good agreement with the real scenario. However, given the behavior of previous states, and the general scenario of Brazil, it is most likely that Sao Paulo founds itself on a 90% loss scenario.

**Fig. 15:**
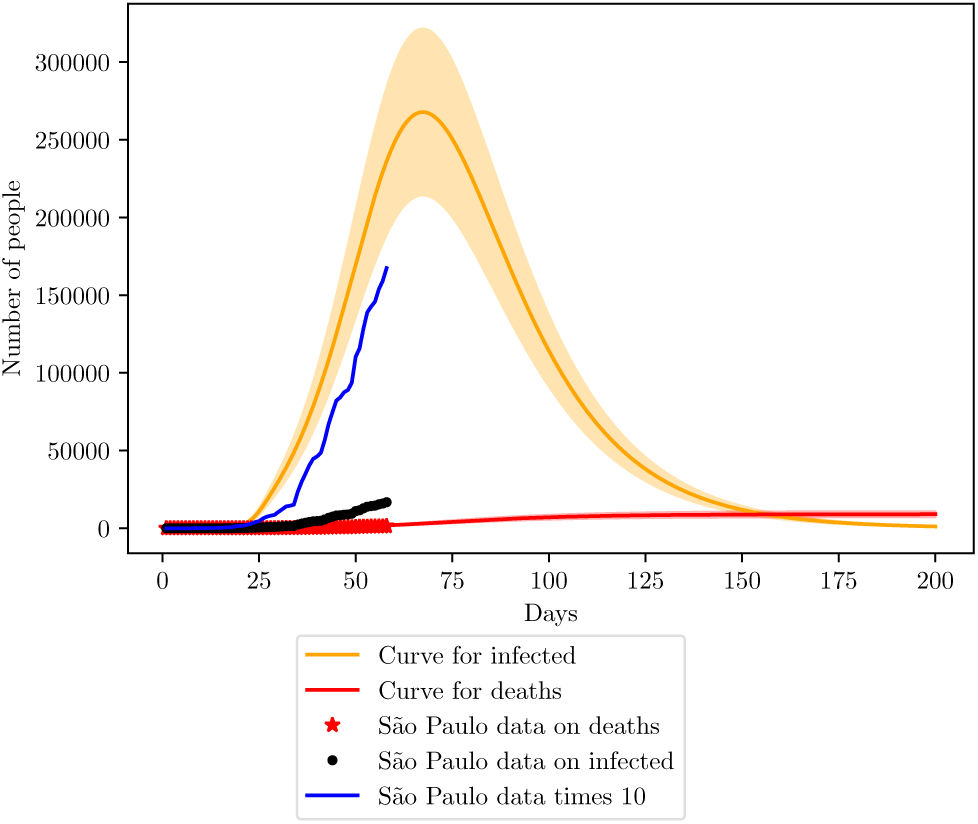
Simulation of the COVID-19 pandemic crisis on Sao Paulo. The blue curve was shown to represent the 90% loss of data on Sao Paulo considering a constant testing rate.

**Fig. 16:**
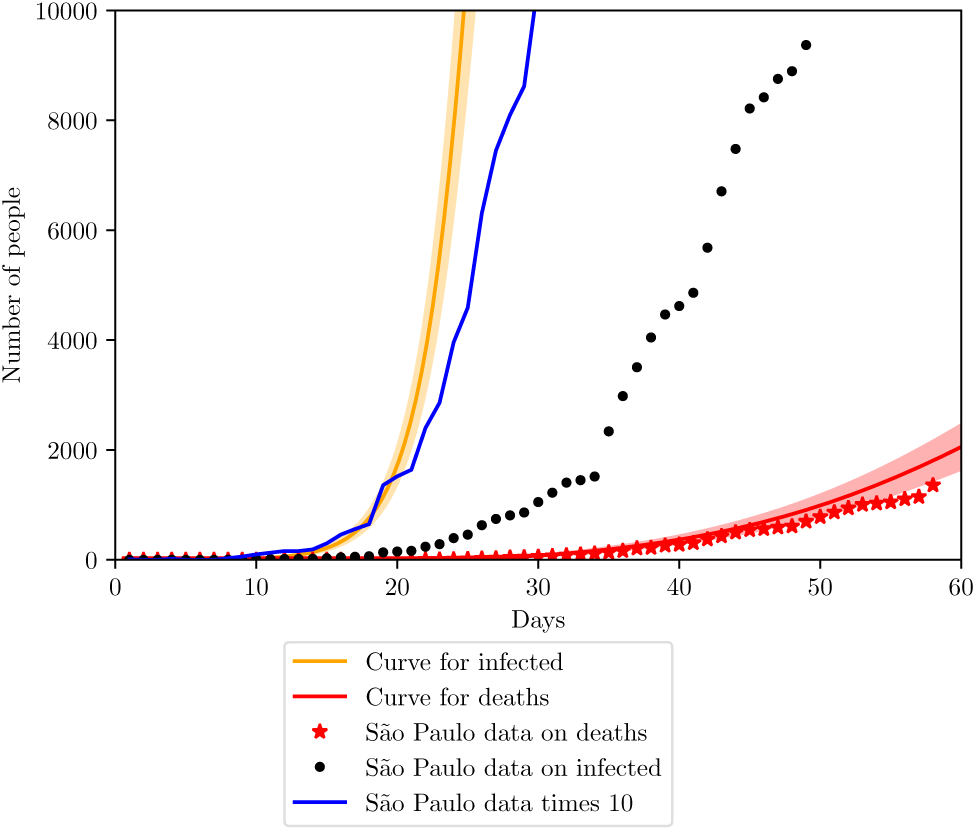
Simulation of the COVID-19 pandemic crisis on Sao Paulo on most recent days.

The state is with it’s peak projected to be around the 70th day of infection, namely close to 7th May. The peak number of infections should be 260000 (208000 - 312000). For the number of deaths, the estimate is close to 6500 (5200 - 7800).

From the AMIB census, the state of Sao Paulo has a total of 7312 ICU beds and recent news point to 53% of them already occupied, leaving around 3400 ICU beds available for COVID-19 treatment.

**Fig. 17:**
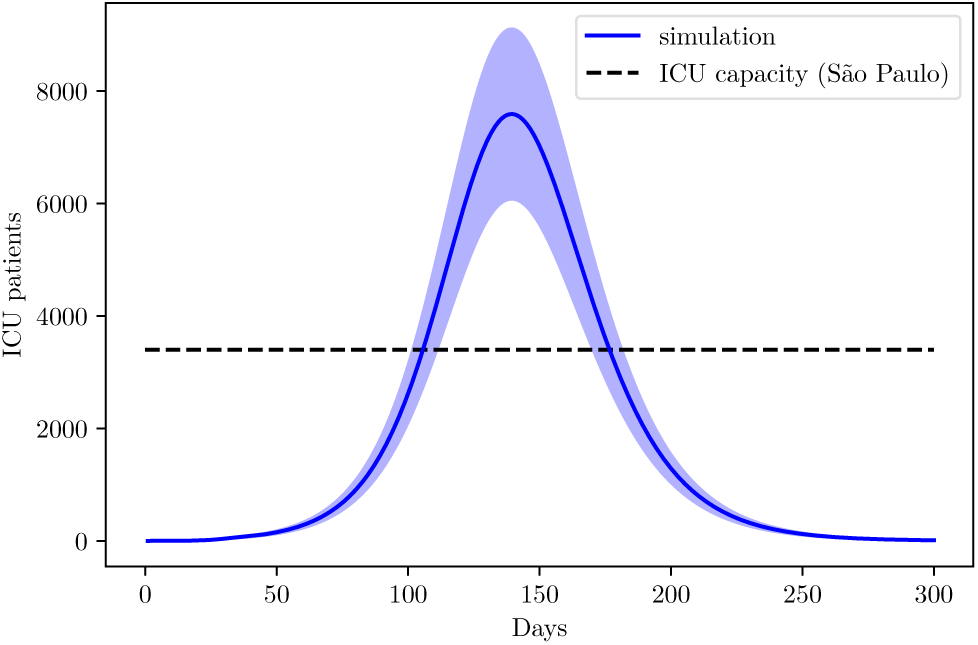
Simulation of the ICU demand on the state of Sao Paulo.

For Amazonas, the fitting of data acquired from the Health Ministry yields *β* = 0.406±0.096 and *τ_d_* = 16± 6, showing that, despite the high number of undocumented infections, the state is on the same situation found on other states. Knowing the behavior of the curve, but not the true number of each point on the curve. The difference from previous states is that the value of *τ_d_* is also in agreement with international values. The average isolation index for Amazonas is around 51%.

The census from IBGE [42] was also used here to acquire population data for the state.

From the AMIB census, Amazonas posses 249 ICU beds, with 55% of them occupied before the outbreak. Unfortunately, no data on tests was found for Amazonas, therefore we consider a 90% loss of infections.

**Fig. 18:**
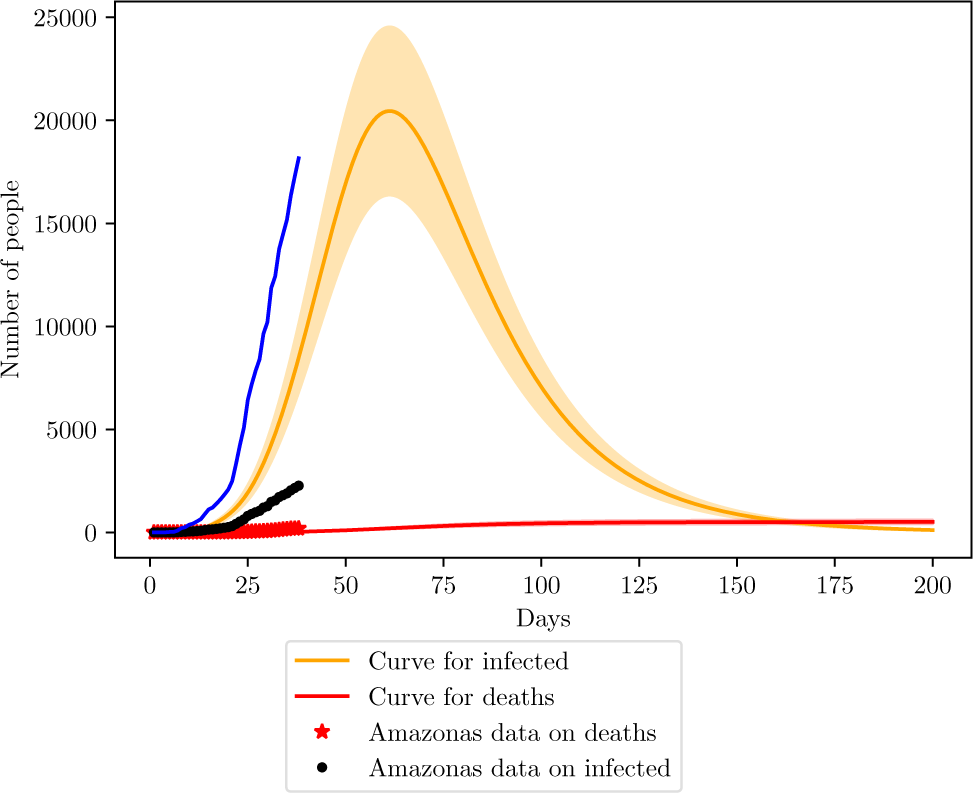
Simulation of the COVID-19 pandemic crisis on Amazonas.

**Fig. 19:**
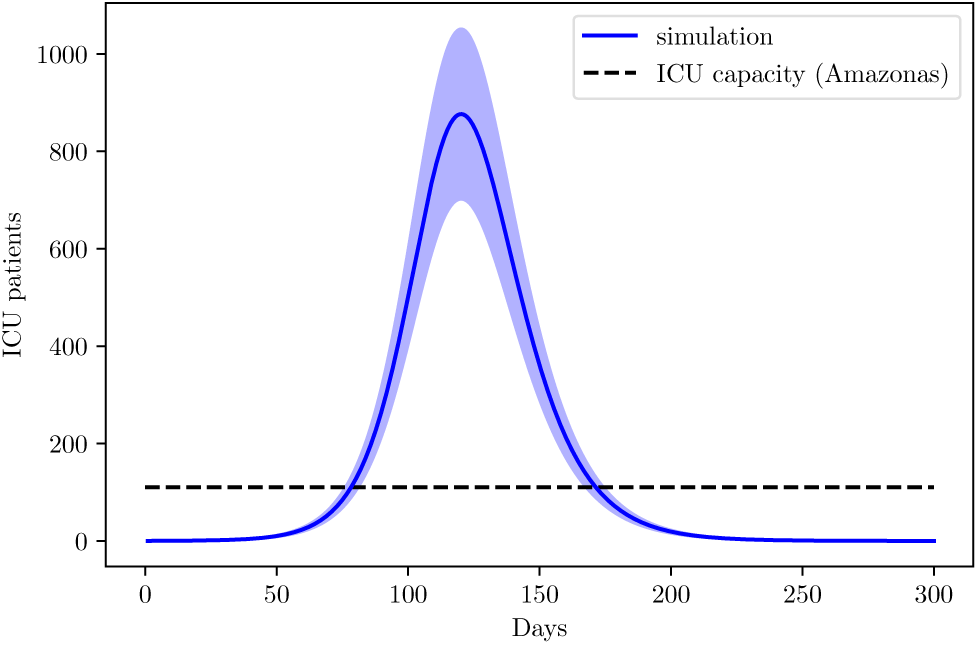
Simulation of the ICU demand on the state of Amazonas.

Amazonas peak is estimated to May 16th, with a total of 20000 infections peak (16000 - 24000). Deaths are estimated to reach 500 (400 - 600).

## VI. FUTURE SCENARIOS

Simulating the stopping of non-pharmaceutical interventions is equivalent to make *β* increase back to it’s starting value. By making such simulations, we observe a increase of cases, that is, a second peak of the disease right after the stop.

**Fig. 20:**
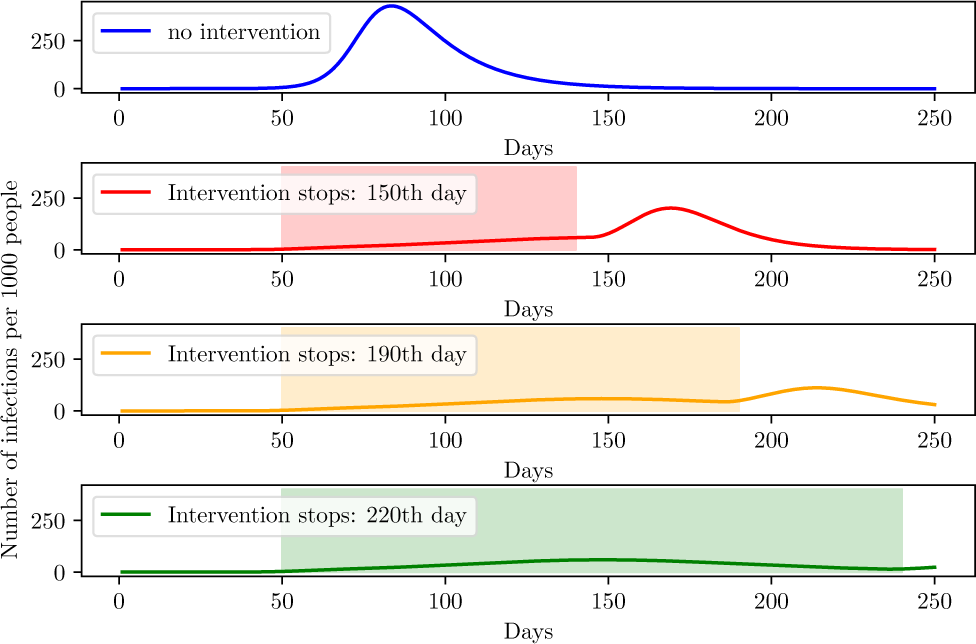
Height of the second peak for different times on social isolation.

Figure 20 shows that to drastically diminish the second peak, the social isolation must endure about 220 days supposing an efficiency of 70%, it is equivalent to state that on Brazil, quarantine should hold until October, while for a total prevention of the second peak, social isolation must take place until December. That is expected and agrees to other simulations made by different groups, another group from the University of Harvard projected that, to prevent a second peak on the world the possible re-incidence of the virus, social isolation must hold until the beginning of 2021 and social distancing until 2022 or 2024 [43].

However, that scenario might drastically change with the introduction of vaccines or efficient medicine on the population. As show on simulations, such pharmaceutical interventions are able to decrease rapidly the infection curve. In order to simulate the effect of medicine on the population, we started decreasing the death probability *P*_:(_ and time taken from symptoms onset to recovery *τ_r_* from a specific date, until it reaches a maximum value. We supposed that the introduction of medicine decreased both *P*_:(_ and *τ_r_* by half on the period of 10 days after the introduction on the population.

For the vaccines, we added the term −*vS*(*t*) on (1), which takes out individuals from the susceptible group at a rate v called vaccination rate, and added the term *vS*(*t*) on (4), adding those individuals on the recovery group, granting them immunity against the virus. The vaccination rate *v* was chosen to behave according to a logistic function starting on 0, and gradually increasing to 0.2 after an specific time.

**Fig. 21:**
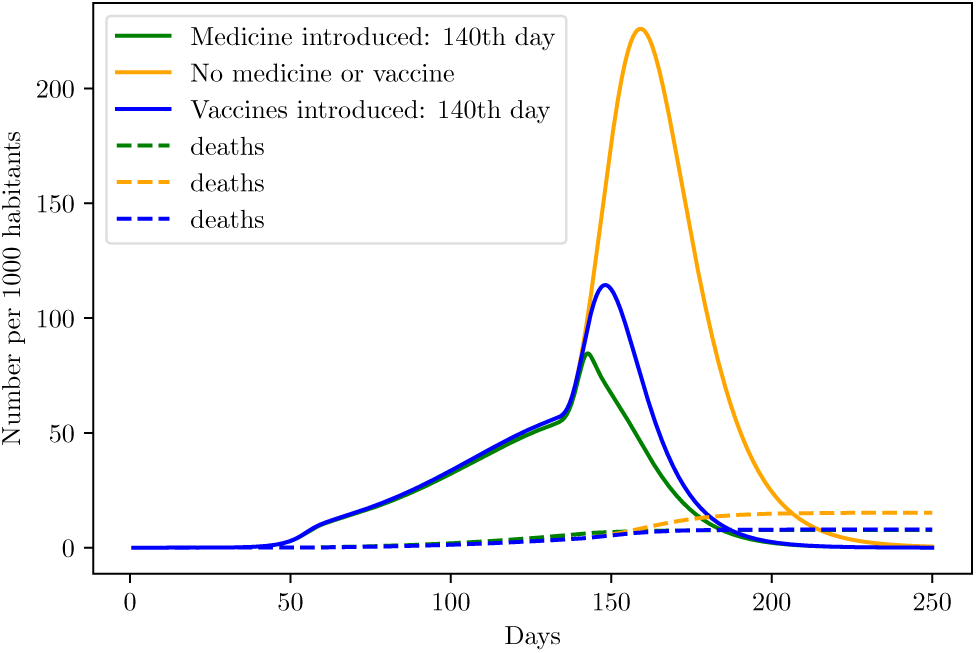
Behavior of the infection curve if the vaccination/medication occurs at the same time of intervention stopping.

**Fig. 22:**
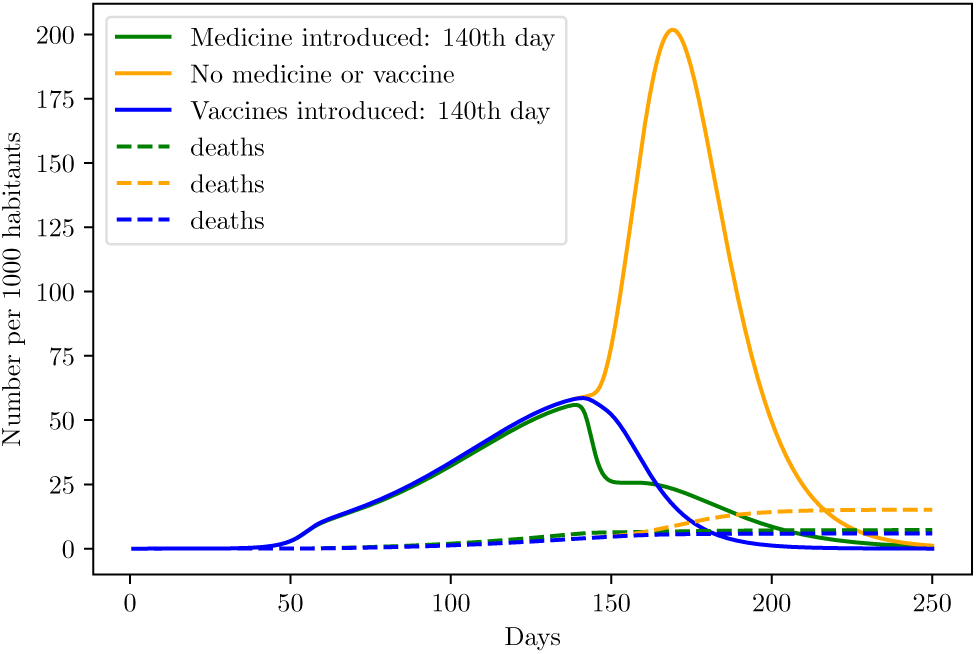
Behavior of the infection curve if the vaccination/medication occurs 10 days before time of intervention stopping.

From the simulations, the most safe method is not to stop the intervention and introduce the vaccines or drugs into the population, but to wait a small period of 10 days before stopping the intervention.

## VII. CONCLUSION

Simulations of the COVID-19 outbreak vary from model to model, here we try to find balance on the most precise model, which could be achieved considering also a group of asymptomatic infections and hospitalized, and the availability of data. In doing so, we decided to simulate the behavior of the disease on Brazil based on international parameters under the assumption that the virus would not be much different on Brazil and the main aspects regarding the transmission would be intervention efficiency, population demographics and social contact. This assumption might prove limited if later should be found that climate effects strongly alters the spread.

Another limitation of the model is on the assumption of homogeneous population. We tried here to counter-attack this limitation by estimating the effective population *N* according to international parameters and by widening the error margin of the predictions. A better estimate of the outbreak could be done by assessing cities individually, however that would represent a loss of data, since demographics available by IBGE regard mainly states and major cities. Another outtake would be the testing data, the states which provided testing data, did only for the whole state but not for individual cities. We did not consider comorbidities on the population such as diabetes and cancer, however the age of the individual seems to be the most important factor on determining mortality factors [44].

We also state here that the nature of the process is stochastic, allowing fluctuations from the deterministic model used to run the simulations. Thus, this study present an estimate of the real situation and expected behavior given the parameters associated with the disease and the efficiency of the intervention. The above results present the dimension of the real scenario, but due to possible initial fluctuations on the stochastic behavior of reality, we might find some deviations from the expectancy.

Even with limitations, the model has proven efficient on generating curves that agree with the estimated loss of cases for each state. From the states studied here, Sao Paulo, Amazonas and Pernambuco present the highest risk of collapse on the health system, while Espirito Santo and Distrito Federal should have minor issues with system collapse or none at all. The blue curve representing the behavior of the official data considering the error percentage for Amazonas exhibited a growth far from the simulation region, however, it falls perfectly inside this region when data is translated by 10 days, meaning that if the infection on Amazonas begun 10 days earlier than previously thought, data fits the simulated curve.

On the duration of social isolation, the safer situation is to hold the isolation for as long as possible in order to decrease the second peak height, while increasing the number of tests performed. All simulations considered here did not assume the end of the intervention, therefore, numbers of deaths may be higher. Should any efficient drugs on combating the virus come along, the simulations shows the safer way is to first introduce them on the population without breaking the social isolation, and about 10 days later start the process of reopening.

## Data Availability

Source code for running a few SEIRD models and SEIRD models with age division are provided on Github/PedroHPCintra.

https://github.com/PedroHPCintra/Coronavirus/blob/master/Coronavirus.ipynb

## VIII. SUPPLEMENTARY MATERIAL

Source code used for some simulations and with didatic example of predictions available at https://github.com/PedroHPCintra/Coronavirus.

